# Rasch validation of a new scale to measure dependency in arm use in daily life: the Upper Limb Lucerne ICF-based Multidisciplinary Observation Scale (UL-LIMOS)

**DOI:** 10.1101/2023.01.26.23285068

**Authors:** Ann Van de Winckel, Beatrice Ottiger, Janne Marieke Veerbeek, Thomas Nyffeler, Tim Vanbellingen

**Author notes:** **Correspondence:** Corresponding Author: Ann Van de Winckel, PhD, MSPT, PT.

## Abstract

**Introduction:** About 77% of adults with stroke have upper limb impairments. Many upper limb measures are available for adults with stroke to measure the impairment and activity level of the affected limb. However, an observational scale focused on assessing dependency in upper limb use during daily life activities (as opposed to testing in laboratory settings) is lacking. To bridge this gap, we have developed a new 5-item “Upper Limb Lucerne ICF-based Multidisciplinary Observation Scale (UL-LIMOS)”, which assesses dependency on others during affected arm use in daily life in adults with stroke. As a next step in the psychometric analysis, we evaluated the unidimensionality and structural validity of the UL-LIMOS with Rasch Measurement Theory.

**Methods:** This is a single-center cross-sectional study in adults with (sub)acute stroke. We applied Rasch Measurement Theory (RMT) to analyze the structural validation and unidimensionality of the new UL-LIMOS. We chose a polytomous partial credit model using the Rasch Unidimensional Measurement Model (RUMM) 2030 software. The outputs provide evidence of unidimensionality, item and person fit, overall fit, principal component analysis of residuals (PCAR), person separation reliability (PSR), as well as residual item correlations to identify local item dependence. Person mean location, floor and ceiling effects identify proper targeting.

**Results:** We recruited 407 adults with (sub)acute stroke (median age 63 years, 157 women). All items and persons fit the Rasch model, and the PSR of 0.90 indicates that clinicians and researchers can reliably use the scale for individual decision-making. There were small floor (2.70%) and ceiling (13.00%) effects. The average person mean location was 1.32 ± 2.99 logits, indicating that the items were too easy for this group of adults with (sub)acute stroke. The PCAR’s eigenvalue was 2.46 with 49.23% explained variance. Further analysis of paired *t*-tests revealed that 0.89% of person locations were significantly different when comparing the two subtests formed based on positive and negative loadings on the first principal component, thereby confirming the unidimensionality of the scale. One pair of items related to “arm and hand use” and “fine hand use” showed residual item correlations.

**Discussion:** The new Rasch-based UL-LIMOS is a valid ICF-based observation scale at the ICF-participation level, to evaluate dependency during upper limb use in daily life in adults with stroke. The UL-LIMOS would be a valuable addition to the core assessments of adults with (sub)acute stroke in hospitals and rehabilitation centers. Further analysis is needed to generalize our findings to adults with chronic stroke who have returned to their home setting, and in other countries to account for cultural differences. Targeting could be improved in the future. Additional psychometric analyses, such as sensitivity to change, are warranted. A comparison of the UL-LIMOS data with self-reported measurements or accelerometers could potentially lead to changes to the core datasets recommended for the evaluation of adults with stroke.

## 1 Introduction

About 77% of adults with stroke have upper limb impairments (1). These impairments hinder performing activities of daily life (ADL) independently and result in long-term dependency in 50% of the cases (2,3). This dependency decreases the quality of life (QoL) (4) and results in an inability to return to work in 40% of working-age adults who have had a stroke (5).

Many upper limb measures evaluate motor recovery after stroke (for an overview see (6)). Following the International Classification of Functioning, Disability and Health (ICF) (7), these measures assess either upper limb impairment (e.g., mobility of joints, muscle power/tone/endurance), or assess activities (e.g., grasping a block of wood) in laboratory settings.

Of all the upper limb measures, the Upper Extremity Subscale of the Fugl-Meyer Motor Assessment (FMA-UE) (8) on the impairment level and the Action Research Arm Test (ARAT) (9) on the activity level were suggested as core upper limb assessments for stroke rehabilitation trials (10) and clinical rehabilitation (11). So far, there is no consensus on scale recommendations for assessing dependency in upper limb use in daily life (10). The Barthel Index (BI) (12) and the Functional Independence Measure (FIM) (13) –originally designed to evaluate the need for nursing care– are commonly used in clinical rehabilitation and research to assess daily life functioning in general (11,14), such as feeding, bathing, dressing, and undressing. However, BI (15) and FIM (16) do not focus on specific upper limb use in daily life. Moreover, problematic floor and ceiling effects (>15%) have been reported (15–17). Some patient-reported outcome measures, such as the Motor Activity Log (MAL) (18), or ABILHAND (19), which are administered through semi-structured interviews, were developed to evaluate the stroke individual’s perspective on real-life upper limb performance. Yet, due to the subjective nature of patients’ reports, these measures should not be used with adults with stroke who have moderate to severe cognitive deficits. For this reason, they were not suggested as core measures to assess upper limb performance in daily life (10). The Actual Amount of Use Test (AAUT) is an observational test, which measures how much patients spontaneously use their affected arm during predefined tasks in a laboratory setting. Therefore, the AAUT also does not reflect spontaneous upper limb use in daily life (20).

Others have used accelerometers as a measure of upper limb performance in daily life, which has the advantage of not being biased by patients’ subjective reports (21,22). However, accelerometers cannot determine what type of activity was performed. In sum, an observational scale specifically focused on assessing dependency in upper limb use during actual daily life activities (as opposed to testing in a laboratory setting) is lacking.

Recently, we demonstrated that the Lucerne ICF-based Multidisciplinary Observation Scale (LIMOS), a clinician-reported measure, was reliable and valid in evaluating the performance of activities in daily life in adults with acute and subacute stroke (23–26). Clinician-reported measures are measures scored by a health care professional based on observing the patients’ spontaneous behaviors, e.g., during their stay in the hospital or rehabilitation center. It can therefore be used in adults with moderate to severe cognitive impairments after stroke. LIMOS covered several domains (motor, communication, learning and applying knowledge, and domestic life), and showed no problematic floor or ceiling effects (23,26). Moreover, responsiveness was higher for LIMOS than for BI and FIM (25).

Based on this previous work, and to fill the gap concerning assessing actual arm and hand use in daily life after a stroke, we developed a new, 5-item upper Limb Lucerne ICF-based Multidisciplinary Observation Scale (UL-LIMOS), which evaluates upper limb use in daily life. The goal of developing this new measure, derived from the reliable and valid LIMOS (23–26), is to obtain a quick evaluation measurement of dependency on others for upper limb use in daily life, for use in the hospital and rehabilitation centers, or in research (27). As a next step in establishing the psychometrics of this scale, we aim to test the structural validity and unidimensionality of the UL-LIMOS, using Rasch measurement Theory.

## 2 Materials and methods

### 2.1 Participants

We approached adults with (sub)acute stroke for participation in the study, who were admitted for inpatient neurorehabilitation in the rehabilitation center Neurocenter, Luzerner Kantonsspital, Lucerne, in Switzerland. Stroke diagnosis was based on the European Stroke Organization (ESO) guidelines, which are based on both clinical and Magnetic Resonance Imaging (MRI) criteria. We included adults with a first-ever acute to subacute stroke, up to 6 months post-stroke, showing unilateral ischemic or hemorrhagic supratentorial lesions (28). Adults with stroke were excluded if they had bilateral lesions. There were no other inclusion and exclusion criteria.

The study was conducted in accordance with the principles of the Declaration of Helsinki (2013) and was approved by the local Ethical Committees of the state of Luzern (BASEC-ID 2017-00998). We followed the Strengthening the Reporting of Observational Studies in Epidemiology (STROBE) Statement (29). The participants gave written informed consent. A family member with power of attorney consented if participants had severe cognitive impairments, preventing them to consent independently.

### 2.2 Main outcome measures

We acquired demographic (sex, age) and clinical data (type and time after stroke, location of stroke, presence of cognitive deficits (30–32), and more specifically, unilateral neglect (33) and apraxia (34–36) for their impact on motor function) as well as the UL-LIMOS at admission. The UL-LIMOS is composed of 5 items, which are items selected from the more encompassing LIMOS (23–26). The LIMOS addressed the dependency of others during daily activities on several domains, among which motor activities (with 18 items).

Our previous reliability and validity studies on LIMOS demonstrated high internal consistency (coefficient α=0.98), good test-retest reliability at the item level (moderate to excellent range of kappa between 0.41 and 0.84, except for two items with fair agreement, kappa = 0.32–0.37), and subscale levels (intraclass correlation coefficient *r*>0.75, range 0.76–0.95) (23,26). Inter-rater reliability demonstrated moderate to excellent agreement with kappa values ranging from 0.41 to 0.92 (23,26) except for 12 items demonstrating fair agreement.

We demonstrated a strong convergent validity between LIMOS motor and FIM motor (*r* = 0.89), between LIMOS motor and Barthel Index (*r* = 0.92), between LIMOS motor and FIM mobility (*r* = 0.90), and between the subscales LIMOS knowledge and FIM cognition (*r* = 0.81) (23,26). Correlations between other subscales of the LIMOS (self-care, general tasks, domestic life) and the subscales of the FIM ranged between *r* = 0.36– 0.79 (23,26). A moderate positive correlation was found between LIMOS cognition and communication subscale and FIM cognition (*r* = 0.67) (23,25,26).

The LIMOS motor subscale, and the applying knowledge and communication subscale were more responsive, expressed by higher effect sizes (ES = 0.65, Standardized Response Mean, SRM = 1.17 and ES = 0.52, SRM = 1.17, respectively) as compared with FIM motor (ES = 0.54, SRM = 0.96) and FIM cognition (ES = 0.41, SRM = 0.88) or Barthel (ES = 0.41, SRM = 0.65) (25).

Rasch-based LIMOS subscales fit the Rasch model after reducing and rescoring items: LIMOS subscales motor (18 items), communication (5 items), applying knowledge/cognition (13 items), and domestic life (5 items) (24).

Regarding **UL-LIMOS**, the 5 items, used in previous studies(27,37)are “*lifting and carrying objects*” (item 1), “*fine hand use*” (item 2), “*hand and arm use*” (item 3), “*washing oneself*” (item 4), and “*dressing*” (item 5). The items were scored on a 5-point scale (0 to 4) with 0 being “patient is not able to fulfill a task or needs assistance up to 75% (corresponding to “complete”)”; 1 representing “patient is able to fulfill tasks with assistance of 25% to 75% (corresponding to “severe”); 2 being “patient is able to fulfill tasks with assistance less than 25% or under supervision (corresponding to “moderate”)”; 3 representing “patient is able to fulfill tasks independently but needs more time and/or with auxiliary materials, aids (corresponding to “slight”)”; and 4 being “patient is able to fulfill tasks independently (corresponding to “none”)”. The 5 items are summed to obtain a total UL-LIMOS score, ranging from 0 representing no use of the upper arm in daily life to 20 representing independent use of the upper limb in daily life (for the manual containing the scoring sheet and instructions, see ***Supplementary Material***).

We previously demonstrated a strong positive correlation (*r*=0.78) between UL-LIMOS and handgrip strength (assessed with the Jamar dynamometer), and a moderate negative correlation (*r*=0.69) with the Catherine Bergego Scale, which quantifies the influence of spatial neglect-related deficits on the ADL (37).

### 2.3 Statistical analysis

The Rasch Measurement Theory (RMT) was applied to analyze the structural validation and unidimensionality of the new UL-LIMOS. We chose a polytomous partial credit model using the Rasch Unidimensional Measurement Model (RUMM) 2030 software.

A major advantage of RMT is that UL-LIMOS items can be hierarchically ordered from easy to difficult and that ordinal scales are converted to interval scales where person and item distributions are located on the same ruler with logit units. The Rasch prediction model states that a person with a higher level of independence in upper limb use in daily life has a higher probability of scoring higher on items than a person with less independence in upper limb use in daily life.

We followed the RULER guideline recommendations for reporting Rasch-based studies, recently published by Van de Winckel et al. (2022); Mallinson et al. (2022) (38,39). In short, to test these assumptions of the prediction model and aspects of unidimensionality, Chi-square statistics are calculated to evaluate the item and person fit, as well as the overall fit of the scale (38,39). The principal component analysis of residuals (PCAR) provides additional information in relation to the unidimensionality of the scale (38–40) with eigenvalues and percentage variance accounted for by each principal component. Further analysis of paired *t*-tests provides evidence that the scale is unidimensional, if less than 5% significant difference is found when comparing the two subtests based on positive and negative loadings on the first principal component of the PCAR.

The person separation reliability (PSR) identifies the measurement’s precision and indicates whether we can reliably separate high from low person ability at a group (PSR ≥ 0.70) or individual level (PSR > 0.90) (38,39,41). Targeting is identified by floor and ceiling effects (>15% considered as problematic), as well as the person mean location relative to the item location, which is by default positioned on the logit scale at 0 logits ± 1 standard deviation (38,39). The scale is well targeted when the person mean location is within 0.5 logits of the item mean (38,39). We identify local item dependence through residual correlations (42). A correlation of at least 0.2 above the average residual item correlation indicates that this pair of items have more in common with each other than with the whole scale (42).

## 3 Results

We recruited 407 adults with (sub)acute stroke (63.2 ± 16.0 years of age; 157 women). The demographic and clinical details are presented in ***Table 1***. All patients were admitted to the rehabilitation center (Neurocenter, Luzerner Kantonsspital, Lucerne, Switzerland) for inpatient neurorehabilitation between January 2014 and November 2016 (24).

**Table 1.**
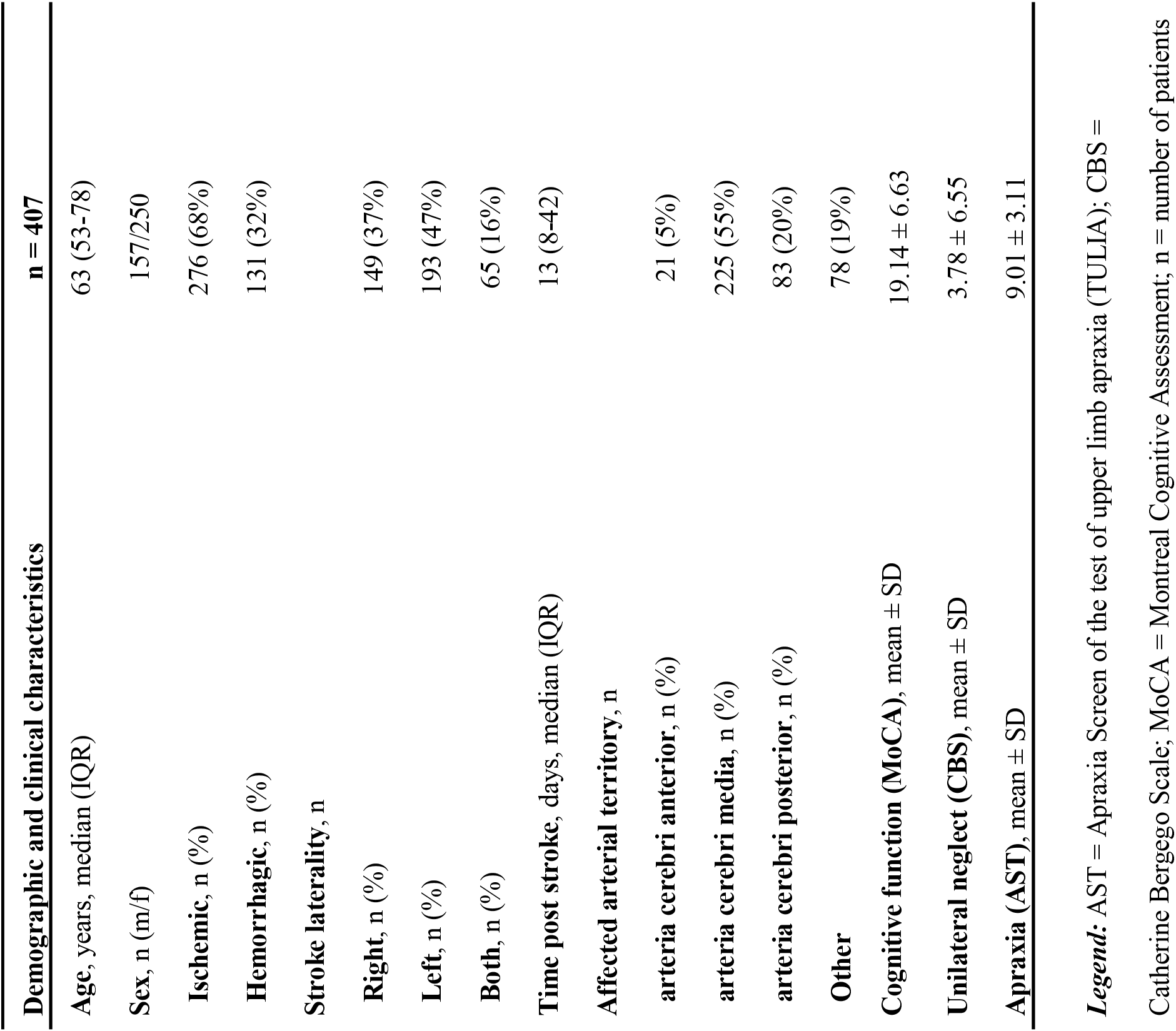
Demographic and clinical characteristics of adults with (sub)acute stroke.

### 3.1 Rasch-based UL-LIMOS

The overall fit, item and person fit, PSR, floor-and ceiling percentages, and PCAR are shown in ***Table 2***. All items and persons fit the model. The UL-LIMOS fit the Rasch model without the need to remove or rescore items. The individual item fit is displayed in ***Table 3***. The threshold map for the Rasch-converted UL-LIMOS is displayed in ***Figure 1A***. This threshold map can be used in the hospital or rehabilitation center for individual patient assessment. The person-item threshold distribution is shown in ***Figure 1B***. The total score of Rasch UL-LIMOS is displayed in ***Table 4***, with the conversion from the original ordinal scores (0 to 20 points) to logits, and logits further converted to a 0 to 100 scoring.

**Table 2.**
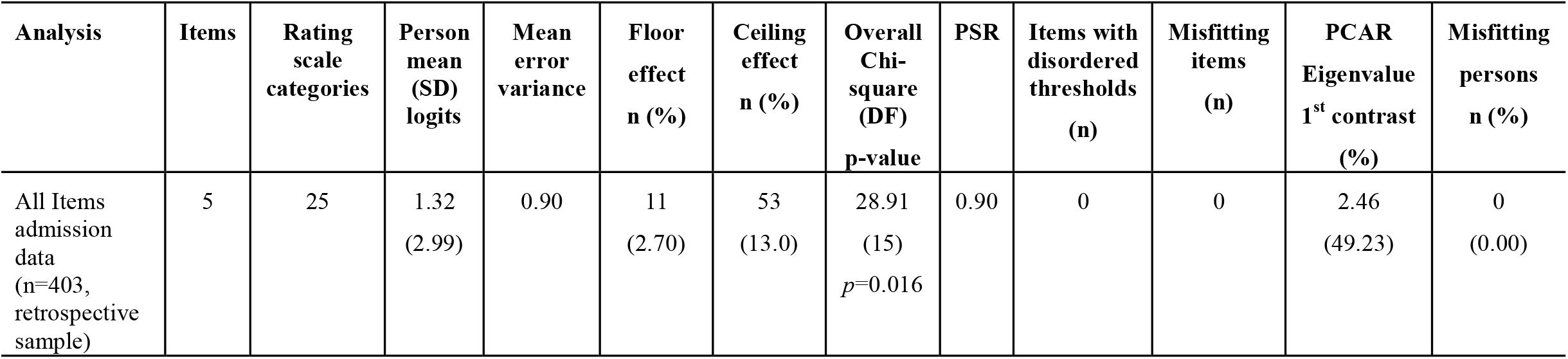
The overall fit, item and person fit, and person separation reliability (PSR), floor-and ceiling values and percentages, and principal component analysis of residuals (PCAR)

| Analysis | Items | Rating<br>scale<br>categories | Person<br>mean<br>(SD)<br>logits | Mean<br>error<br>variance | Floor<br>effect<br>n (%) | Ceiling<br>effect<br>n (%) | Overall<br>Chi-square<br>(DF)<br>p-value | PSR | Items with<br>disordered<br>thresholds<br>(n) | Misfitting<br>items<br>(n) | PCAR<br>Eigenvalue<br>1<sup>st</sup> contrast<br>(%) | Misfitting<br>persons<br>n (%) |
| --- | --- | --- | --- | --- | --- | --- | --- | --- | --- | --- | --- | --- |
| All Items<br>admission<br>data<br>(n=403,<br>retrospective<br>sample) | 5 | 25 | 1.32<br>(2.99) | 0.90 | 11<br>(2.70) | 53<br>(13.0) | 28.91<br>(15)<br>*p*=0.016 | 0.90 | 0 | 0 | 2.46<br>(49.23) | 0<br>(0.00) |

**Table 3.** Item fit statistics of the Rasch-based UL-LIMOS scale.

| Item number | Location<br>(logits) | SE | Chi Square | Item Fit<br>Residual | *p*-value |
| --- | --- | --- | --- | --- | --- |
| UL-LIMOS |  |  |  |  |  |
| 1. Hand and arm use | -0.56 | 0.09 | 3.33 | -1.60 | 0.34 |
| 2. Fine hand use | -0.19 | 0.09 | 2.24 | -0.97 | 0.52 |
| 3. Washing oneself | 0.12 | 0.09 | 8.95 | -2.05 | 0.03 |
| 4. Lifting and carrying objects | 0.29 | 0.08 | 6.11 | 6.32 | 0.10 |
| 5. Dressing | 0.33 | 0.08 | 8.07 | -2.82 | 0.04 |

Legend: SE: Standard Error; *p*-values are Bonferroni corrected for multiple comparisons ( $\alpha=0.01$ for 5 items)

**Table 4.** Converted scoring system for the total UL-LIMOS score.

| **Raw score** | **logits** | **Converted logits to 0-100** |
| --- | --- | --- |
| **UL-LIMOS** |  |  |
| 0 | -6.38 | 0 |
| 1 | -5.17 | 10 |
| 2 | -4.09 | 19 |
| 3 | -3.21 | 27 |
| 4 | -2.37 | 34 |
| 5 | -1.79 | 39 |
| 6 | -1.33 | 43 |
| 7 | -0.92 | 46 |
| 8 | -0.56 | 49 |
| 9 | -0.22 | 52 |
| 10 | 0.11 | 55 |
| 11 | 0.44 | 58 |
| 12 | 0.79 | 61 |
| 13 | 1.16 | 64 |
| 14 | 1.57 | 67 |
| 15 | 2.04 | 71 |
| 16 | 2.56 | 76 |
| 17 | 3.12 | 80 |
| 18 | 3.74 | 86 |
| 19 | 4.50 | 92 |
| 20 | 5.45 | 100 |

**Figure 1.**
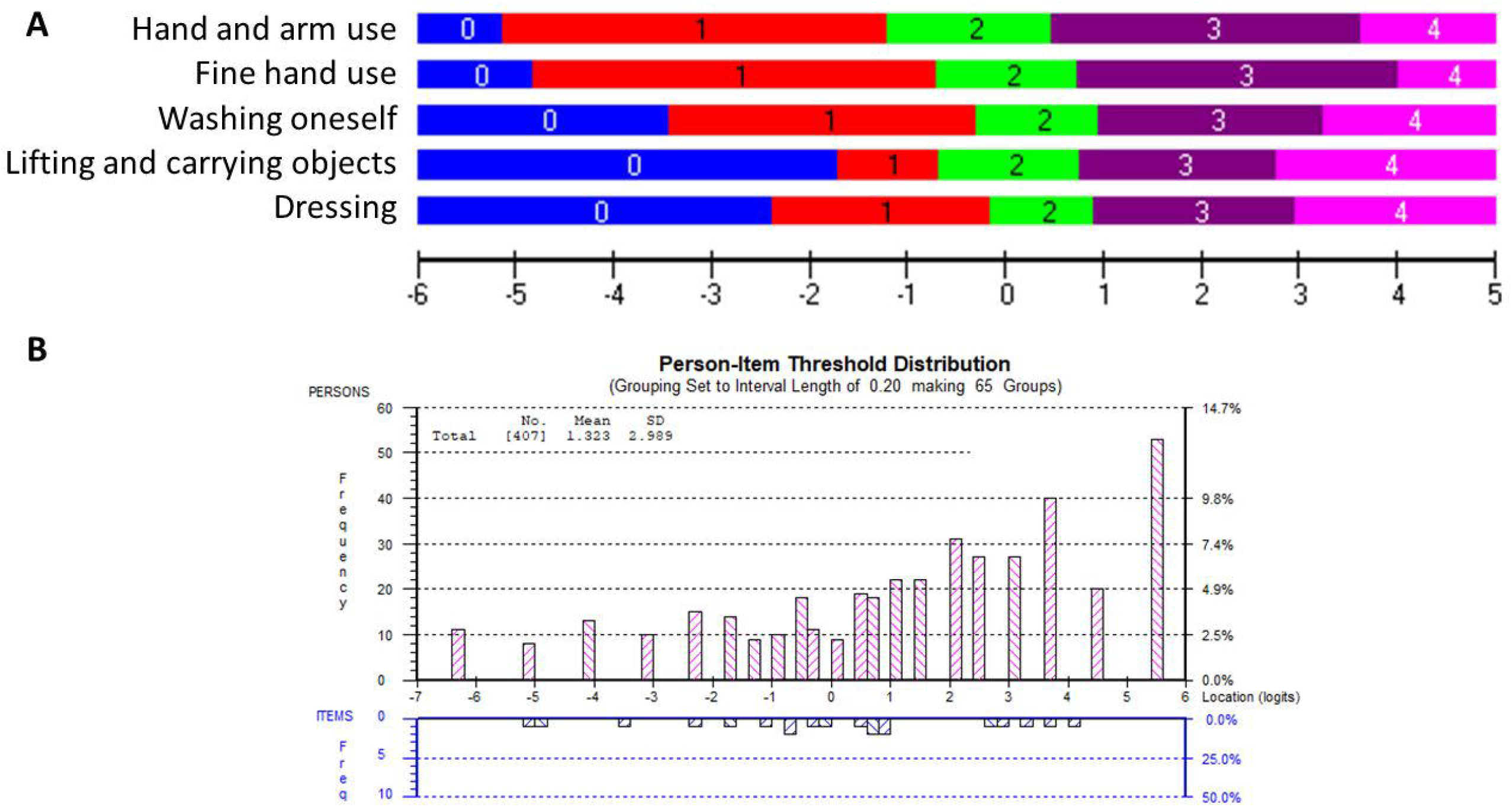
**A. The Rasch-based UL-LIMOS scale: Item threshold map:** The item threshold map depicts the difficulty of the items from the easiest item at the top to the most difficult item at the bottom along with the scoring categories. These item thresholds are matched on the same logit scale (horizontal black line at the bottom of the picture) as the person’s ability. This is a visual depiction of the interval scale, using the same color coding for each item threshold. This demonstrates that with increasing ability, it is easier to get a higher score on an easy item than on a difficult item. It also shows what score would be expected for each item, based on a person’s ability. **B. The Rasch-based UL-LIMOS scale: Person-item threshold distribution:** The ability of the persons (top, pink bars) is plotted on the same logit scale as the difficulty of the item thresholds (bottom, blue bars). The histograms depict the frequency of persons at a certain ability level, from a low ability on the left to a high ability on the right side of the ruler. The number of item thresholds is organized in increasing difficulty levels from the easiest on the left to the most difficult item thresholds on the right side of the ruler.

Around 13.00% of participants obtained a maximum score, meaning there was a small but not problematic ceiling effect. There was also a small (2.70%) floor effect. The average person mean location was 1.32 ± 2.99 logits, indicating that the items were too easy for this group of adults with (sub)acute stroke. PSR was 0.90, meaning the scale can reliably distinguish individuals of different ability levels for decision-making in research and in the hospital or rehabilitation center (39).

The PCAR’s eigenvalue on the first contrast was 2.46 with 49.23% explained variance on the first principal component. Further analysis of paired *t*-tests revealed that 0.89% of person locations are significantly different when comparing the two subtests formed based on positive (items 1 and 2) and negative loadings (items 3 and 5) on the first principal component, thereby confirming the unidimensionality of the scale.

Only one pair of items (items 1 “*lifting and carrying objects*” and item 2 “*fine hand use*”; *r* = 0.65) was above 0.2 of the average residual item correlations (*r* = 0.43). Both specifically identify hand use, which could explain why they are more strongly related to each other than to the rest of the items.

## 4 Discussion

This study presents a new valid ICF-based observation scale (UL-LIMOS) to evaluate dependency during upper limb use in daily life in adults with stroke at the ICF-participation level. Structural validity of the UL-LIMOS was evaluated with Rasch analysis in 407 adults with (sub)acute stroke, demonstrating that UL-LIMOS fit the model, without problematic floor or ceiling effects, and with a high PSR of 0.90, which allows clinicians and researchers to use the scale for individual decision-making.

Rasch analysis also provided insight into the hierarchy of difficulty of the five items. As expected, based on the conceptual framework of upper limb movements, fine hand use requires more dependency than arm and hand use. Tasks such as washing and dressing are even more difficult because they require more awareness and interaction with the whole body, and/or require more cognitive motor planning regarding the different motor action sequences to perform the activity. This is reflected within the Rasch-based hierarchical order of the items.

Dressing appeared to be the most difficult item, confirming previous descriptions of dressing as a complex skill that requires several physical motor function skills and cognitive abilities (43). Notably, approximately 50% of adults with stroke still cannot dress independently six months post-stroke (44). Cognitive deficits could be an important factor for this dependency, and, among the spectrum of different cognitive factors, spatial neglect has been shown to have a major negative impact on dressing skills in adults with a right hemispheric stroke (43,45). Upper limb apraxia has been shown to affect dressing in adults with a left hemispheric stroke (38,39). In our sample, 78% had deficits in cognition (30–32), and, more specifically, 39% had spatial neglect (33), and 35% had moderate to severe apraxia (34–36), further confirming the problematic factors, influencing motor actions that were previously identified in the literature. We confirm earlier findings in the literature that more adults with right hemispheric stroke exhibit neglect and more adults with a left hemispheric stroke have upper limb apraxia. However, the influence of these cognitive disorders on the use of the upper limb in daily life needs to be studied, and assessed comprehensively, in much more detail in future studies.

Evaluating upper limb motor impairment and/or activity in a structured, laboratory-based setting (‘capacity’), such as Fugl-Meyer (8), MESUPES (46), and ARAT (9), and comparing those results to their performance level with UL-LIMOS, measuring reliance on others for upper limb use in daily life, is important because these outcomes may not always line up. Adults with stroke may have the ability to recruit motor units to perform specific motor actions in a laboratory setting but may not be able to generate the necessary motor programs or have the necessary cognitive processing skills to perform tasks in a more unstructured and more complex environment such as is the case with daily life activities.

Evaluation scales are often used in the hospital and rehabilitation settings to provide some estimates to patients regarding their recovery potential and which treatments would be most appropriate for them. Therefore, early prediction algorithms have gained much attraction in recent years (47–51). Yet, the current upper limb prediction models test upper limb motor function in a structured laboratory setting,(47) which does not reflect actual upper limb use in daily life. In addition, these studies often exclude adults with cognitive deficits post-stroke (48–50,52). Thus, the predictions are only applicable to a subset of adults with stroke given the high prevalence of cognitive deficits after stroke, including spatial neglect (53) and apraxia (35). Interestingly, Stinear et al. (2017), who developed the prediction algorithms PREP and PREP-2, emphasized the importance of including cognitive factors in future prediction paradigms, because these factors influenced upper limb outcomes (48,51). This stresses the need for new predictive models that consider the evaluation of dependency on others during upper limb use in daily life. The UL-LIMOS, which describes dependency on others during spontaneous upper limb use in daily life, can also be used in patients with neglect and apraxia. Therefore, UL-LIMOS, as well as measures of neglect and apraxia could therefore be valuable factors in future predictive models of upper limb motor recovery after stroke.

Our study has limitations. Adults with stroke were recruited in only one neurorehabilitation center in Switzerland, which could limit the generalizability to other countries with different cultures. Furthermore, targeting could be improved in the future by adding more difficult items to the scale. Other psychometrics such as sensitivity to change need to be performed on the 5-item UL-LIMOS. Lastly, the scale also needs to be validated in adults with chronic stroke to ensure the generalizability of the results.

In conclusion, we present a new 5-item Rasch-based UL-LIMOS scale, which was validated in 407 adults with acute or subacute stroke. We recommend validation of the UL-LIMOS in other countries to account for cultural differences. The UL-LIMOS could also be validated in chronic stroke stages when adults have returned to their home setting. A comparison of the UL-LIMOS data with self-reported measurements or with accelerometers could potentially lead to changes to the existing core datasets recommended for the evaluation of upper limb performance of adults with stroke (10,11).

## Supporting information

MANUAL UL-LIMOS

## Data Availability

Original datasets requests can be directed to the first or last author.

## 6 Author Contributions

TV, BO, JV, and TN contributed to the conception and design of the study. TV organized the database. AVDW performed the statistical analysis and the interpretation of the results, and she wrote the first draft of the manuscript. TV wrote sections of the manuscript. All authors contributed to the manuscript revision. They also read and approved the submitted version.

## 8 Acknowledgments

We thank the neurorehabilitation team involved in the data collection and all participants for their time and investment. We like to express our deep gratitude to Marc Noël for the critical review of the manuscript.

## 9 Data Availability Statement

Original datasets requests can be directed to the first or last author.

## 10 Contribution to the field statement

Many adults with stroke experience problems using their affected arm in daily life. Until today, however, there is no measure that evaluates how much adults with stroke need to rely on others when using the affected arm in daily life.

To bridge this important gap, we have developed a new 5-item “Upper Limb Lucerne ICF-based Multidisciplinary Observation Scale (UL-LIMOS)”, which reliably assesses dependency on others during affected arm use in daily life in adults with stroke. Our study examining 407 adults with (sub)acute stroke shows that questions in the UL-LIMOS are valid and are measuring what they are supposed to measure. The UL-LIMOS could be used in new predictive models of upper limb motor recovery after stroke because of the demonstrated structural validity without problematic floor or ceiling effects. Our results also show that clinicians and researchers can use the scale for individual decision-making.

