## Supplementary material for "Rasch validation of a new scale to measure dependency in arm use in daily life: the Upper Limb Lucerne ICF-based Multidisciplinary Observation Scale (UL-LIMOS)": MANUAL UL-LIMOS

### 1 Supplementary Data

Number and Title of each element:

Page 2 to 4: Introduction UL-LIMOS Manual

Page 5: Case Report Form

Pages 6 to 10: Instructions UL-LIMOS

Upper Limb - Lucerne ICF-Based Multidisciplinary Observation Scale

(UL-LIMOS) Manual

The UL-LIMOS is an observation based on the International Classification of Functioning, Disability, and Health (ICF), to evaluate upper limb (hand/arm) use in real-life settings. It is a clinician-reported outcome where health professionals are rating patients with stroke based on their observation of the patient’s behaviors in the clinic or rehabilitation center. This can be done in the first 72 hours after admission to neurorehabilitation, then weekly, and in the last 72 hours before discharge from inpatient and/or outpatient neurorehabilitation.

The assessment of items is evaluated on the basis of the assistance that the patient needs to use his/her upper limb (hand/arm) in real-life settings. Item selection was based on ICF Health Brief Core Set for Stroke (Geyh *et al.* 2004) and other literature (Quintas *et al.* 2012).

The criteria for each of the ratings on the 5-point scale are as follows:

0	=	patient is not able to fulfill a task or needs assistance up to 75% (corresponding to “com-

plete”)

1	=	patient is able to fulfill tasks with assistance of 25% to 75% (corresponding to “severe”)

2	=	patient is able to fulfill tasks with assistance less than 25% or under supervision (corre-	sponding to “moderate”)

3	=	patient is able to fulfill tasks independently but needs more time and/or with auxiliary mate-

rials, aids (corresponding to “slight”)

4	=	patient is able to fulfill tasks independently (corresponding to “none”)

**CASE REPORT FORM**

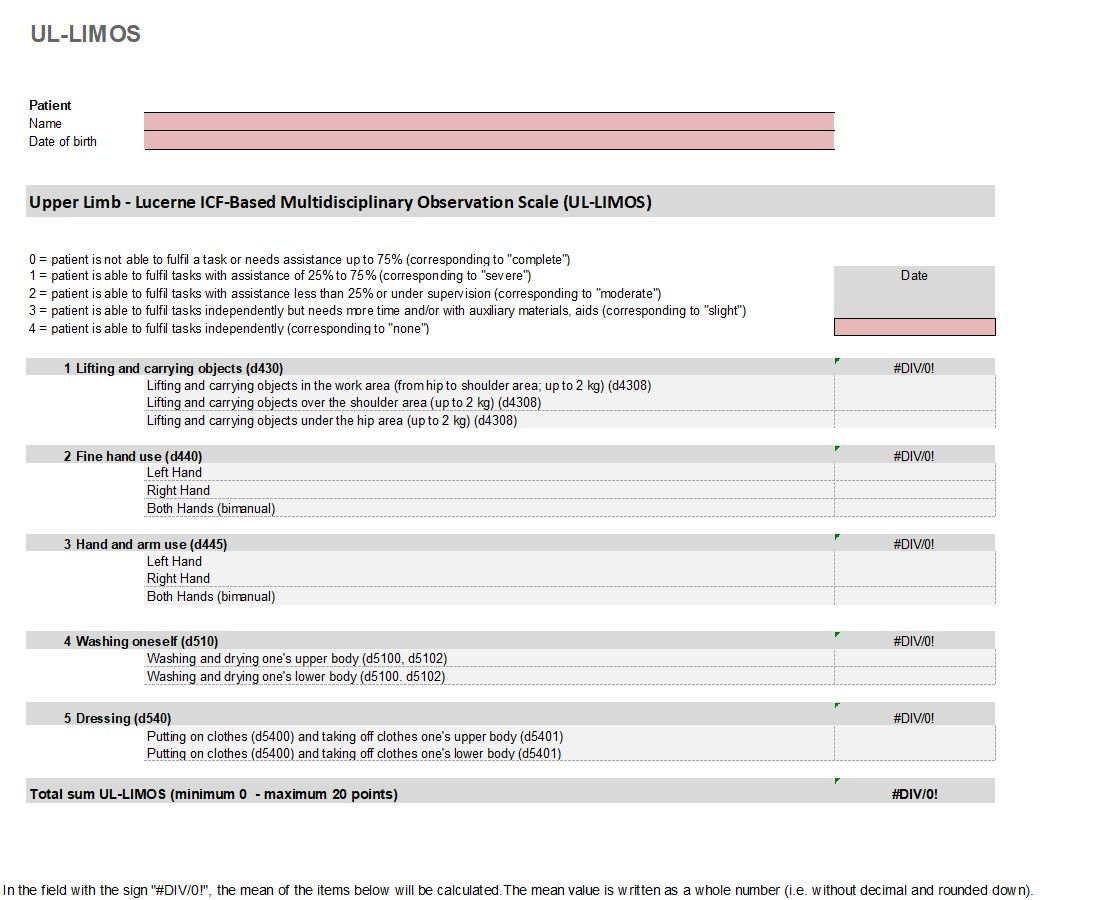

**INSTRUCTIONS**

All items need to be observed. The clinician will not ask the participant to perform specific tasks.

Below is more detailed information on how to score each item. In addition, we recommend every user to do a 45 minutes workshop to learn how to use the UL-LIMOS in clinical practice.

### 1 Lifting and carrying objects (d430)

| ICF Definition | Raising up an object or taking something from one place to another, such as when lifting a cup or carrying a child from one room to another.  Inclusions: lifting, carrying in the hands or arms, or on shoulders, hip, back or head; putting down. |
| --- | --- |
| Preamble | This item is assessed regardless of whether the patient walks or uses a wheelchair. This item is assessed separately for the work area (from hip to shoulder area) (d4308), over the shoulder area (d4308), as well as for under the hip area (d4308). The weight of the objects is up to 2 kg. |
| **Legend** | **Examples** |
| **0** | - The patient contributes less than 25% to movement execution. - The patient is unable to lift and carry an item (e.g., a ball, balance pad, pillow) (overhead; below the hips). |
| **1** | - The patient initiates movements or performs partial steps of the movement sequence (assistance of 25% to 75%). - The patient needs physical/structural help to lift and carry an item. |
| **2** | - The patient requires less than 25% assistance (verbal/tactile) in executing the movement, e.g., because the patient drops the object or cannot maintain balance. - For safety reasons, the patient needs supervision. |
| **3** | - The patient takes longer to lift and transport an item. |
| **4** | - The patient lifts objects independently and transports them. |

### 2 Fine hand use (d440)

| ICF Definition | Performing the coordinated actions of handling objects, picking up, manipulating, and releasing them using one's hand, fingers, and thumb, such as required to lift coins off a table or turn a dial or knob.  Inclusions: picking up, grasping, manipulating, and releasing. |
| --- | --- |
| Preamble | This item is assessed separately for the left and right hand, as well as for bimanual hand use. Examples for fine hand use are buttoning and unbuttoning shirt buttons, tying shoelaces, removing coins from purse, etc |
| **Legend** | **Examples** |
| **0** | - The in-hand manipulation of objects is not possible and must be fully compensated. |
| **1** | - Fine motor movements are not used efficiently on objects. The patient requires tactile guidance (assistance of 25% to 75%) from an assistant. - For example, the patient keeps dropping the objects. |
| **2** | - The object can be held in the hand, the patient can execute pinch grips. - Use of assistance of less than 25% (e.g., support surfaces, tactile guidance) or aids are still needed, or the movements are uncoordinated. |
| **3** | - In-hand manipulation of objects is possible, but the patient takes longer to execute the task. - Autonomous in-hand manipulation of objects is possible with aids. |
| **4** | - In-hand manipulation of objects is performed skilfully. |

### 3 Hand and arm use (d445)

| ICF Definition | Performing the coordinated actions required to move objects or to manipulate them by using hands and arms, such as when turning door handles or throwing or catching an object.  Inclusions: pulling or pushing objects; reaching; turning or twisting the hands or arms; throwing; catching. |
| --- | --- |
| Preamble | Hand use is observed during gross motor activities. Fine motor hand skills are observed in Item 2. This item is assessed separately for the left and right hand, as well as for bimanual hand and arm use. |
| **Legend** | **Examples** |
| **0** | - The patient cannot use his/her hands and arms in everyday life and needs maximum assistance to perform gross motor activities. |
| **1** | - The patient needs moderate to significant tactile guidance (assistance of 25% to 75%) and/or a significantly adjusted setting (e.g., positioning, decrease in gravity) to enable hand and arm use in daily life. |
| **2** | - The patient only needs tactile guidance for selected movements (less than 25% assistance), such as for movements over 90 ° flexion/abduction, in order to be able to use hands and arms in daily life. |
| **3** | - The patient performs movements with hands and arms autonomously, takes longer to execute the task and/or needs aids in daily life (e.g., tongs). |
| **4** | - The patient uses hands and arms in daily life in a coordinated and skilful manner. |

### 4 Washing oneself (d510)

| ICF  Definition | Washing and drying one's whole body or body parts, using water and appropriate cleaning and drying materials or methods, such as bathing, showering, washing hands and feet, face, and hair, and drying with a towel. |
| --- | --- |
| Preamble | This item is assessed separately for washing and drying one’s upper body (d5100, d5102) and one’s lower body (d5100, d5102). Place of execution is not relevant. The shower and bathing tools (bath towels, shower gel) are available. For hygiene reasons, catheter care in patients with indwelling catheter is always performed by the nursing staff. |
| **Legend** | **Examples** |
| **0** | - The patient contributes less than 25% to washing and drying of his/her upper/lower body. - All steps of the action must be guided and structured. - The patient does not want to wash his/her upper body. |
| **1** | - The patient initiates or executes individual steps (assistance of 25% to 75%). - The patient needs physical/structural support to carry out the procedure. - The patient begins to wash himself/herself only after verbal and/or tactile input from an assistant. |
| **2** | - The patient performs several steps of the procedure (assistance of up to 25%, and supervision). - The patient needs brief tactile and/or verbal inputs to continue/ complete the action. - The patient needs supervision during the activity for safety reasons. |
| **3** | - The patient washes and dries his/her upper/lower body independently but takes longer to execute the task. - The patient washes and dries his/her upper/lower body independently but needs aids. The patient needs to be checked upon once, whilst washing and drying his/her upper body. |
| **4** | - The patient washes and dries his/her upper/lower body independently within a normal time frame. |

### 5 Dressing (d540)

| ICF  Definition | Carrying out the coordinated actions and tasks of putting on and taking off clothing and footwear in sequence and in keeping with climatic and social conditions, such as by putting on, adjusting, and removing shirts, skirts, blouses, pants, undergarments, saris, kimono, tights, hats, gloves, coats, shoes, boots, sandals, and slippers. |
| --- | --- |
| Preamble | This item is assessed separately for putting on clothes (d5400) and taking off clothes (d5401) one’s upper and lower body. |
| **Legend** | **Examples** |
| **0** | - The patient contributes less than 25% to the action. - All steps of the action must be guided and structured. |
| **1** | - The patient initiates or executes individual steps (assistance of 25% to 75%). - The patient needs physical/structural support to carry out the procedure. - The patient puts on clothes only after verbal and/or tactile input from an assistant. |
| **2** | - The patient performs several steps of the procedure (assistance of up to 25%, and supervision). - The patient needs brief tactile and/or verbal inputs to continue or complete the action (e.g., buttoning/unbuttoning, zipping/unzipping). - The patient needs supervision for safety reasons. |
| **3** | - The patient dresses his/her upper/lower body independently but needs to use an aid or appliance to be able to dress, and/or takes longer to execute the task. - The patient needs to be checked upon once. |
| **4** | - The patient dresses his/her upper/lower body independently within a normal time frame. |
